# Association of Adduction-Induced Optic Nerve Head Strain with Visual Field Loss Progression in Myopic Normal-Tension Glaucoma *The Wenzhou Glaucoma Progression Study*

**DOI:** 10.64898/2026.10.15.26365686

**Authors:** De-Fu Chen, Zhuxin Xiong, Jinrui Zhang, Chunfang Deng, Jia Li, Guohui Liu, Tao Wang, Qiangjie Huang, Jost B Jonas, Michael J A Girard, Xiaofei Wang, Yuanbo Liang

## Abstract

**Purpose:** To characterize the biomechanical response of the optic nerve head (ONH) in myopic versus non-myopic individuals with normal-tension glaucoma (NTG), and to evaluate the association between ONH strains and the rate of visual field (VF) progression.

**Design:** Retrospective clinical cohort study.

**Participants:** A total of 118 NTG eyes from 77 patients with ≥ 2 years of follow-up and 5 VF tests were enrolled, including 66 eyes with myopia and 52 eyes without.

**Methods:** Each participant underwent three-dimensional (3D) optical coherence tomography (OCT) imaging of the ONH under three mechanical loading conditions: 20° adduction, 30° abduction and acute intraocular pressure (IOP) elevation by applying a 0.80 N force to the lower eyelid. Effective strains across the global ONH complex and within specific segmented tissues (pre-lamina, lamina cribrosa, choroid, and sclera) were quantified using a validated 3D strain-mapping algorithm.

**Results:** Myopic NTG eyes exhibited significantly higher ONH strains compared to non-myopic eyes under acute IOP elevation (10.1±2.7% vs. 9.3±1.8%, P = 0.044) and abduction (9.2±2.3% vs. 8.4±2.1%, P = 0.048). In the non-myopic group, ONH strains varied significantly across conditions (P < 0.001), with adduction (9.9± 2.0%) and acute IOP elevation (9.3±1.8%) eliciting greater strains than abduction (8.4±2.1%). These condition-dependent variations were less pronounced in the myopic group (10.1±2.7% vs. 9.8±2.7% vs. 9.2±2.3%, P = 0.07). Multivariable linear mixed-effects analysis revealed that in myopic NTG, higher ONH strains induced by adduction were significantly associated with faster MD decline (estimate: -0.139 dB/year per 1% increase, P = 0.03, adjusted for age, spherical equivalent and baseline perimetric mean defect). Conversely, larger strains during acute IOP elevation were associated with slower progression (estimate: 0.184 dB/year per 1% increase, P = 0.004). Tissue-specific analysis confirmed that adduction-induced strains in the choroid and pre-lamina tissue as well as IOP-induced strains across all ONH tissues were significantly correlated with the rate of perimetric progression in myopic eyes (all P < 0.05).

**Conclusions:** In this cohort, greater adduction-induced ONH strain was associated with faster perimetric deterioration in myopic NTG. These findings support further investigation of gaze-related optic nerve traction as a potential non-IOP-related biomechanical factor in myopic NTG.

## INTRODUCTION

Glaucoma is a chronic and progressive optic neuropathy, characterized by the loss of retinal ganglion cells (RGCs) due to an axonal damage occurring within the lamina cribrosa (LC).^[1, 2]^ It has been recognized that forces such as those generated by the intraocular pressure (IOP) and orbital cerebrospinal fluid pressure (CSFP), deform the LC as the critical anatomical element in the pathogenesis of glaucomatous optic neuropathy.^[3]^ Previous population-based studies have revealed that normal-tension glaucoma (NTG), with the IOP in the statistically normal range, is more prevalent in East Asia than in Western countries.^[4–6]^ Other studies reported that 30% to 50% of patients with NTG showed a disease progression over a five-year follow-up period, despite IOP levels in the normal range.^[7–9]^ Recent studies by Girard, Demer and others indicated that besides the forces exerted by the IOP and orbital CSFP, other biomechanical forces such as those generated by an optic nerve backward pull during eye movements in axially elongated eyes may also lead to substantial deformations of the ONH.^[10–12]^

Myopia has long been recognized as a risk factor for glaucoma,^[13–15]^ the prevalence of which is increasing globally and is projected to affect approximately 50% of the world’s population by 2050.^[16]^ Recent findings suggest that biomechanical characteristics within the eye, particularly those associated with myopia, may increase the biomechanical vulnerability of the ONH, thereby elevating the risk of glaucoma.^[17–19]^ This association is particularly relevant for NTG, in which elevated IOP is thought to play a smaller role than in high-IOP glaucoma.^[20–22]^ Despite these advances, it has remained unclear how these biomechanical factors influence the progression of NTG. Additionally, differences in ONH strain characteristics between myopic and emmetropic NTG patients may reveal potential mechanistic distinctions, if any, between the two glaucoma subgroups.

This retrospective longitudinal analysis sought to evaluate how the biomechanical responses of the ONH to variations in IOP elevation and gaze changes correlate with the progression of glaucomatous visual field (VF) loss in NTG patients with myopia.

## METHODS

### Study Participants Recruitment

Patients identified with NTG were recruited from the Wenzhou Glaucoma Progression Study (WGPS), a prospective cohort investigation. This research complied with the ethical guidelines set forth in the Declaration of Helsinki and obtained authorization from the Ethics Committee of the Eye Hospital of Wenzhou Medical University (No. KYK-2013-12). Each participant provided written informed consent upon enrollment.

A comprehensive description of NTG and the methodology of the WGPS, including the specific inclusion and exclusion criteria, is available in previous publications.^[23–25]^ For the present investigation, we recruited 77 NTG patients from the WGPS according to following conditions in which patients had been subjected to more than five reliable VF tests and had at least a two-year follow-up period. Reliable VF assessments were characterized by a rate of false positives of 15% or less and rate of fixation losses of 33% or less, with no established limits for false negative.^[24]^ Glaucoma progression was determined based on VF progression following the Early Manifest Glaucoma Trial (EMGT) criteria,^[26]^ utilizing the guided progression analysis through the FORUM Glaucoma Workplace software (version 4.2, Carl Zeiss Meditec AG, Jena, Germany). Myopia was classified as a spherical equivalent of -0.50 diopters (D) or less prior to undergoing refractive or cataract surgery.

During the follow-up visit, patients underwent comprehensive eye examinations that comprised: (1) refractometry using the Nidek ARK-510A Autorefractometer (NIDEK Co. Ltd, Tokyo, Japan); (2) IOP measurement by Goldmann applanation tonometry (GAT) (HAAG-STREIT 900 CM; Haag-Streit, Kloniz, Switzerland) and (3) evaluation of axial length and central corneal thickness utilizing the Lenstar 900 (Haag-Streit, Kloniz, Switzerland).

### Visual Field Testing

VF tests were performed every 3 to 6 months using standard automated perimetry (Humphrey750i, Zeiss Corporation, Germany). The Humphrey Field Analyzer employed the central 24-2 protocol based on the Swedish Interactive Threshold Algorithm Standard.^[27]^

### OCT Imaging under Baseline and Mechanical Loading Conditions

Prior to imaging, pupillary dilation was achieved by administering 1.0% tropicamide to each subject (Supplementary Figure). The ONH was imaged using a swept-source optical coherence tomography (SSOCT) system (VG200D; SVision Imaging Ltd., Henan, China) operating in a dense raster scan mode. The system utilizes a laser with a central wavelength of 1,050 nm and operates at a scanning speed of 200,000 A-scans per second. Each volume scan covered a 6 × 6 mm rectangular region centered on the ONH, consisting of 512 serial B-scans with 512 A-scans per B-scan. The mean distance between adjacent B-scans was 11.7 |im, yielding average axial and lateral pixel resolutions of 3.23 |im and 11.7 jim, respectively. A total of four scans were acquired per eye under distinct conditions: a baseline scan in the primary gaze at a baseline IOP level, and three scans under mechanical loading conditions (20° adduction, 30° abduction, and acute IOP elevation).

For the gaze-induced loading conditions, OCT scans were obtained in two specific gaze positions: nasal gaze (20° adduction) and temporal gaze (30° abduction), both without artificial IOP alteration. The eye movements were induced by turning the subject’s head while the patients maintained visual fixation on the stationary OCT objective target. Specifically, for the right eye, 20° adduction was achieved by rotating the head 20° to the right, whereas 30° abduction was achieved by rotating the head 30° to the left, with the eye continuously fixated straight ahead. This directional protocol was reversed for the left eye. To ensure movement precision, the exact angle of head rotation was continuously monitored using an inertial measurement unit (IMU) sensor mounted centrally on the subject’s forehead.

For the IOP-induced loading condition, imaging was performed in the primary gaze while a constant mechanical force of 0.8 N was applied to the temporal aspect of the lower eyelid using an ophthalmodynamometer (ODM), following previously established protocols^^[11, 17, 28]^ Briefly, the ODM was oriented perpendicular to the anterior sclera to gently exert an external force. An eyeglass frame fitted with a short circular tube of spiral steel wire guided the indentation bar to ensure consistent and uniform direction across subjects. This application of force was maintained consistently throughout the OCT scanning procedure. Baseline IOP was measured using GAT prior to pupillary dilation and OCT imaging. Because direct IOP measurement during continuous SSOCT scanning with the ODM in place is technically unfeasible, the magnitude of the acute IOP elevation was estimated based on a prior pilot study involving 40 subjects. In that pilot cohort, IOP was measured using GAT at the slit lamp immediately after the application of the identical 0.8 N force to the lower eyelid. The pilot data confirmed that this controlled mechanical loading acutely elevated the IOP to approximately 35-40 mmHg, which remained stable for the brief duration of the simulated imaging process. This stability was achieved because adequate pupillary dilation and the ultra-fast SSOCT system (200,000 A-scans/sec) enabled each scan to be completed in just a few seconds, thereby avoiding any significant tonographic effect.

### Image Enhancement with Adaptive Compensation

To enhance the visualization of optic nerve head (ONH) structures, particularly the lamina cribrosa (LC), the raw optical coherence tomography (OCT) images were processed using adaptive compensation. As an advanced OCT post-processing technique, adaptive compensation effectively mitigates blood vessel shadowing artifacts and augments tissue contrast. It has been shown to improve the visibility of the ONH structures including the anterior part of the LC and its insertions, alongside other relevant ocular structures.^[29, 30]^

### ONH Reconstruction through Automatic Segmentation

Segmentation of the ONH was performed by fine-tuning the medical image segmentation foundation model, SAM-Med2D. Detailed procedures regarding model fine-tuning and the resulting segmentation performance are provided in the Supplementary Materials. For each ONH, the following tissue groups were automatically segmented: the pre-lamina tissue (PLT, including the retina), the choroid, the visible anterior portion of the peripapillary sclera, and the visible anterior portion of the LC. The fully segmented and three-dimensionally reconstructed ONH served a dual purpose: first, it was used to delineate the volumetric mesh domains required for the subsequent 3D Digital Volume Correlation (DVC) algorithm; second, it enabled the aggregation of calculated strains within selected tissue regions for downstream tissue-specific analysis. Furthermore, in addition to the four aforementioned tissue regions designated for strain analysis, Bruch’s membrane was independently segmented. Bruch’s membrane opening (BMO) points were subsequently extracted automatically by identifying the anatomical terminations of Bruch’s membrane. These BMO points define a distinct geometric plane (the BMO plane), which was utilized as a standard horizontal reference plane for each ONH.

### In Vivo Displacement and Strain Mapping of the ONH

The methodology employed was highly consistent with previously reported studies and we further validated this approach for the new OCT machine and imaging protocol. Initially, each OCT volume acquired under a biomechanical load (i.e., adduction, abduction, or elevated IOP) was digitally registered to its corresponding baseline primary gaze volume utilizing commercial software (Amira V.2020.3; Thermo Fisher Scientific, Waltham, MA, USA). This registration step was essential to eliminate confounding rigid-body translations and rotations arising from subject head or eye movements.

To ensure unbiased inter-group comparisons (e.g., myopia versus non-myopia) regarding 3D deformations and strains, the en-face field of view was cropped to a 2.8 × 2.8 mm² region centered on the BMO for all ONHs. Only the image data within this strictly defined spatial domain were utilized for the subsequent digital volume correlation (DVC) computations and downstream analyses. DVC was performed using the XDigitalVolumeCorrelation Extension in Amira using a two-stage local-to-global strategy. First, a subset-based local DVC approach (DVC Local Approach module; isotropic sub-volume size, 30 jim) was used to capture the initial three-dimensional tissue displacements between paired OCT volumes. The resulting displacement field was then used to initialize a finite-element-based global DVC algorithm, implemented using an isotropic hexahedral mesh size of 100 x 100 x 100 |im and a displacement convergence tolerance (3u) of 0.01 |im, to refine the displacement field. The final displacement field obtained from the global DVC analysis was used for subsequent strain calculations. This procedure was applied to three specific OCT volume pairs for each eye: (1) primary gaze at baseline IOP versus acute IOP elevation, (2) primary gaze versus adduction at baseline IOP, and (3) primary gaze versus abduction at baseline IOP. Additional DVC parameters, strain derivation procedures, and validation of DVC accuracy are provided in the Supplementary Materials. For quantitative comparison, “global ONH strain” was defined as the average effective strain calculated across the entire 3D ONH tissue volume within the cropped region. Additionally, tissue-specific strains were obtained by averaging the effective strains within each corresponding segmented tissue layer.

### Statistical Analysis

All statistical analyses and visualizations were conducted using R version 4.1.3 (R Foundation for Statistical Computing, Vienna, Austria). Categorical variables are presented as counts and percentages (n, %), while continuous variables are expressed as mean ± standard deviation (SD). To evaluate the demographic and clinical features between the two glaucoma subgroups while accounting for inter-eye correlation, a hierarchical approach was implemented. Patient-level parameters (e.g., age and gender) were compared using the Student’s t-test or Chi-square. For eye-specific clinical metrics, linear mixed-effects models (LMM) were utilized for continuous variables and generalized estimating equations (GEE) for categorical data. Both models incorporated the subject ID as a random effect or cluster to adjust for the inherent dependency between binocular data.

To further assess the differences in ONH deformations under varying acute IOP elevation and gaze directions, a multi-level LMM framework were employed, followed by the Bonferroni correction for post hoc pairwise comparisons. To identify potential risk factors associated with glaucoma progression, both univariate and multivariable linear mixed models with subject-specific random intercepts were constructed. This approach ensured that the nested structure of eye-level data was appropriately addressed, thereby preventing the inflation of Type I error. Clinical variables demonstrating a trend toward significance (P ≤ 0.20) in univariate assessments were included in the final multivariable models.

## RESULTS

### Demographics and Clinical Characteristics

A total of 118 eyes with NTG from 77 patients were evaluated, including 66 eyes in the myopic group and 52 in the non-myopic group (Table 1). Patients in the myopic group were significantly younger than the non-myopic group (mean age: 54.0 ± 15.0 vs. 66.4 ± 11.3 years, P < 0.001). As expected, the myopic group exhibited longer axial length (25.6 ± 1.5 mm, range from 22.6 to 29.0 mm vs. 23.2 ± 0.6 mm, range from 21.9 to 24.6 mm; P < 0.001) and more negative spherical equivalent (-4.58 ± 2.93 vs. 0.98 ± 0.98 D, both P < 0.001). Baseline clinical parameters, including follow-up duration, central corneal thickness, IOP, and mean perimetric deviation (MD), were comparable between the two groups (all P > 0.05). Notably, the percentage of male was significantly lower in non-myopic patients, compared to the myopic group (28.8% vs. 54.5%, P = 0.009).

**Table 1.** Demographics and Ocular Characteristics in Normal Tension Glaucoma Patients between Myopic and Non-Myopic Group. CCT = central cornea thickness, IOP = intraocular pressure, MD = mean deviation, DH = disc hemorrhage. Significant P values are shown in bold type.

### ONH Effective Strains in Myopic and Non-myopic Group

As illustrated in Figure 1, myopic NTG eyes exhibited significantly greater global ONH strains (averaged across all tissues) than non-myopic under acute IOP elevation (10.1 ± 2.7 vs. 9.3 ± 1.8%, P = 0.04) and 30° abduction (9.2 vs. 8.4%, P = 0.048). In the non-myopic group, ONH strains varied significantly across different conditions (P < 0.001, Table 2), with acute IOP elevation (9.3 ± 1.8%) and 20° adduction (9.9 ± 2.0%) eliciting markedly higher strains than 30° abduction (8.4 ± 2.1%). In contrast, such condition-dependent variations were less pronounced in the myopic group (P = 0.07). Tissue-specific analysis further revealed that in non-myopic eyes, all components (sclera, choroid, PLT, and LC) showed significant strain variations across conditions (all P ≤ 0.005) (Table 3). However, in myopic eyes, significant variations were confined to the PLT (P = 0.04) and LC (P = 0.02), where acute IOP elevation induced higher strains compared to abduction (PLT: 10.6 ± 2.7% vs. 9.6 ± 2.3 %; LC: 11.3 ± 2.9% vs. 10.3 ± 2.8%).

**Figure 1.**
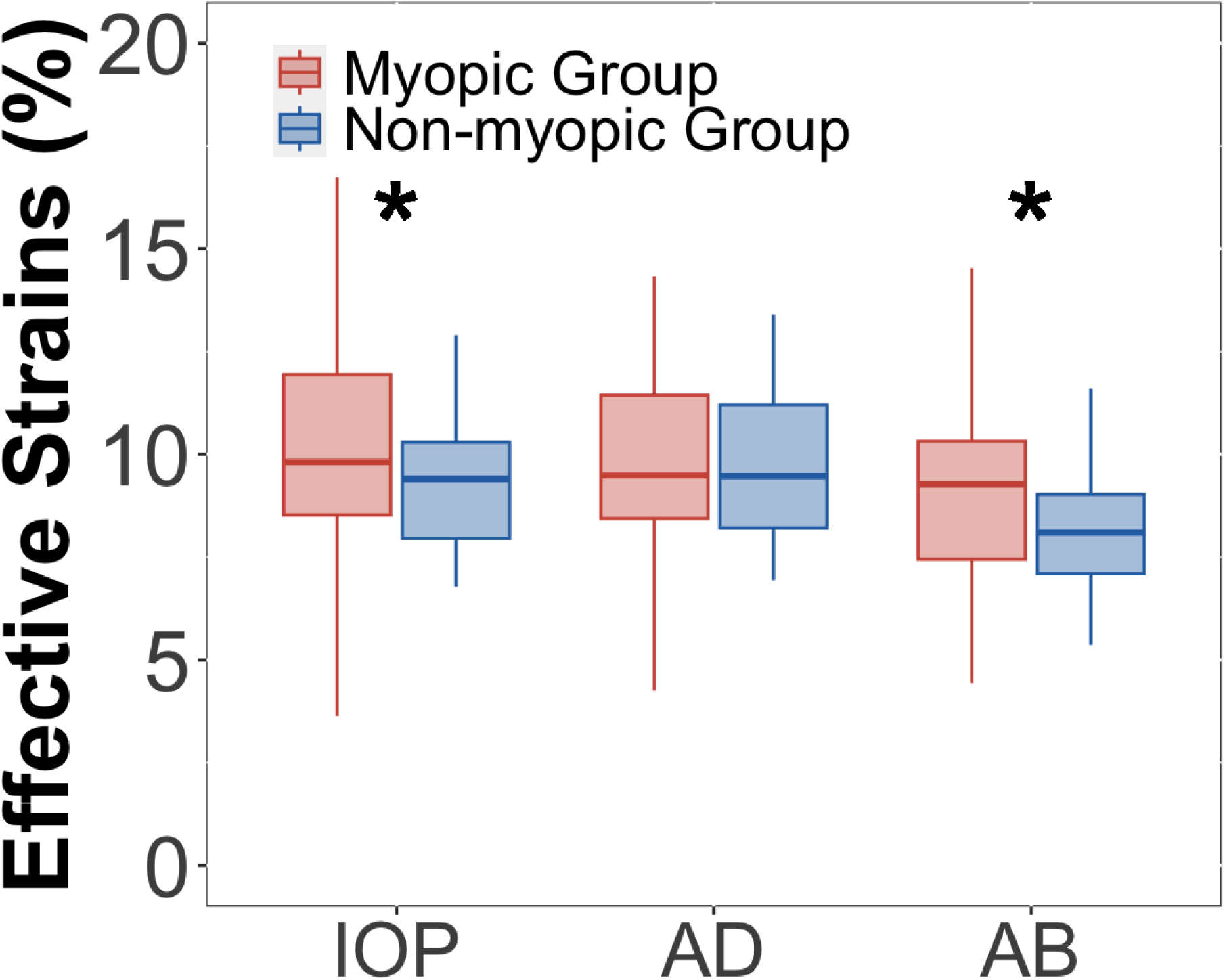
Effective Strains in Myopic and Non-Myopic Group under Various Conditions.

**Table 2.** Effective strains in Myopic and Non-myopic Group. IOP = intraocular pressure, AD = adduction, AB = abduction. Significant P values are shown in bold type. Post hoc significant differences are shown as “>”.

**Table 3.** Effective Strains under Various Conditions in Myopic and Non-myopic Group. IOP = intraocular pressure, AD = adduction, AB = abduction. Significant P values are shown in bold type. Post hoc significant differences are shown as “>”.

### Association Between Effective Strains and Visual Field Progression

The relationship between ONH biomechanics and VF progression was analyzed using linear mixed models (Table 4). In myopic NTG, global strains induced by adduction were significantly associated with accelerated VF progression in the multivariable model (estimate: -0.14 dB/year per 1% increase, P = 0.03). Interestingly, higher global strains during acute IOP elevation appeared to correlate with slower MD decline (estimate: 0.18 dB/year per 1% increase, P = 0.004). Furthermore, severe VF loss at the time of OCT assessment was a strong predictor of faster progression in both myopic and non-myopic groups (both P < 0.001). Tissue-specific modeling (Table 5) highlighted that in the myopic group, adduction-induced strains in the choroid and PLT, as well as acute IOP elevation-induced strains across all tissues (sclera, choroid, PLT, and LC), were significantly correlated with the rate of MD progression (all P < 0.05). In contrast, neither global nor tissue-specific ONH strains under any mechanical loading condition were significantly associated with the rate of visual field progression in the non-myopic NTG cohort (all P > 0.05).

**Table 4.** Linear Mixed-Effects Regression Models Evaluating the Effect of Characteristics and Effective Strains under Various Conditions in the Rates of Mean Deviation Loss Over Time. CCT = central cornea thickness, IOP = intraocular pressure, MD = mean deviation, DH = disc hemorrhage.

**Table 5.** Linear Mixed-Effects Regression Models Evaluating the Effect of Characteristics and Tissue-specific Effective Strains under AD in the Rates of Mean Deviation Loss Over Time. CCT = central cornea thickness, IOP = intraocular pressure, MD = mean deviation, DH = disc hemorrhage.

## DISCUSSION

In the present study, myopic eyes with NTG exhibited significantly greater global ONH deformations compared to non-myopic eyes under various mechanical loading conditions, including acute IOP elevation and abduction. Notably, our multivariable analysis revealed a distinct and contrasting impact of these strains on functional deterioration in the myopic cohort. Adduction-induced strains were significantly associated with accelerated VF progression, whereas greater strains during acute IOP elevation correlated with a slower rate of MD decline. The robust association between adduction-induced strain and functional loss suggests that the mechanical traction exerted during horizontal eye movements may serve as a non-IOP dependent mechanism driving progression of optic nerve damage in myopic NTG patients. Consequently, the pathophysiology of optic nerve damage in the myopic NTG population likely involves an interplay between axial elongation and gaze-induced mechanical shear effects and potentially other factors.

Our findings indicated a strong correlation between adduction-induced strains and the rate of MD decline in the myopic NTG group, a relationship conspicuously absent in the non-myopic group. This observation is intriguing given that both groups exhibited comparable IOP levels and VF defects at the time when the OCTs were taken. Previous work has suggested that the relationship between ONH biomechanics and functional damage may differ between glaucoma subtypes. Chuangsuwanich et al.^[31]^ reported that IOP-induced ONH strain was strongly associated with retinal sensitivity in HTG, whereas this relationship was largely absent in NTG, and proposed that non-IOP-related mechanisms, including optic nerve traction during eye movements, may contribute more prominently to NTG pathophysiology. Our findings extend this concept by showing that, within NTG, the biomechanical response to adduction is associated with longitudinal VF progression specifically in myopic eyes, supporting the possibility that eye-movement-induced loading represents an important additional biomechanical pathway in myopic NTG.

Tissue-specific analyses further showed that the association between adduction-induced strain and VF progression was significant in the choroid and PLT, but not in the peripapillary sclera or LC. This spatial pattern may provide insight into how optic nerve traction is transmitted through the myopic ONH. With increasing myopia, the peripapillary sclera becomes thinner and may exhibit altered mechanical properties, potentially reducing its structural rigidity. During adduction, traction generated by the optic nerve and its sheath is transmitted to the posterior sclera at the optic nerve–scleral junction, probably at the optic nerve dura mater-peripapillary scleral flange junction. A more compliant peripapillary sclera may undergo greater deformation under this load, which may in turn redistribute deformation to adjacent intraocular tissues. Because the PLT and choroid are more compliant than the scleral connective tissue, they may undergo greater local deformation in response to displacement of the surrounding load-bearing structures. Such deformation may be particularly relevant to glaucoma progression because the PLT contains unmyelinated retinal ganglion cell axons, whereas the short posterior ciliary arteries forming the peripapillary arterial circle of Zinn-Haller and located close to the peripapillary choroid provide vascular support to the ONH. Thus, the observed associations in the PLT and choroid may indicate that gaze-induced mechanical loading preferentially affects neural and vascular tissues that are directly relevant to glaucomatous damage.

Interestingly, higher global strains as well as tissue-specific strains across all ONH layers during acute IOP elevation correlated with a slower rate of MD decline in the myopic group in our cohort. While elevated IOP is classically recognized as a primary risk factor for glaucoma progression, our observation suggests that great tissue compliance (reflected as higher strain) might offer a protective buffering capacity specificity against IOP fluctuations in myopic NTG. In the LC, mechanical stresses are dictated by both circumferential traction from the peripapillary sclera (hoop stress) and the anteroposterior trans-LC pressure differential between the IOP and the orbital cerebrospinal fluid pressure (CSFP).^[3, 32]^ While acute IOP elevation inevitably increases both the trans-LC gradient and hoop stress,^[33–35]^ the resulting higher strain observed in these eyes reflects greater tissue compliance.^[35]^ We postulate that this compliant ONH complex acts as a mechanical buffer, by safely deforming under load, the tissue absorbs and dissipates mechanical energy that would otherwise be transmitted directly as destructive stress to the unmyelinated retinal ganglion cell axons. This raises an important biomechanical paradox: why does greater tissue deformability appear protective against acute IOP elevation, yet detrimental during adduction? We hypothesize that this discrepancy stems from the fundamentally different nature of the applied forces. Acute IOP elevation induces a relatively symmetric, expansile stress across the ONH. In this context, a compliant tissue safely yields (acting as a ‘shock-absorber’) to dissipate energy. In contrast, horizontal eye movements like adduction generate asymmetric, tractional shearing forces via the tethered optic nerve. When the ONH complex is overly compliant, it lacks the structural rigidity needed to mechanically resist this backward extraocular pull. Consequently, the tissue undergoes profound local structural distortion (manifesting as excessive strain), leading to direct shearing damage to the delicate unmyelinated axons within the PLT and choroid. Thus, high tissue compliance may be advantageous for buffering internal pressure spikes, but highly deleterious when counteracting asymmetric extraocular traction.

Previous in vivo and computational studies have shown that optic nerve traction during horizontal eye movements can produce substantial ONH deformation.^[36–38]^ The present study revealed that 20° adduction-induced strains throughout the ONH were significantly larger than those induced by 30° abduction only in the non-myopic group. However, Liu et al.^[37]^ reported that adduction induced significantly higher strains than abduction in non-highly myopic (non-HM) eyes, a cohort with a spherical equivalent similar to that of the myopic group in our study (-4.58 ± 2.93 D vs. -5.74 ± 2.40 D). Furthermore, Liu et al. found a positive correlation between higher effective strains and longer axial length, with a more pronounced difference in effective strains induced by adduction (P = 0.002) than by abduction (P = 0.016) between HM and non-HM eyes. In our study, however, adduction-induced effective strains were not significantly higher than abduction-induced strains in the myopic group. These discrepancies may partly be attributable to the limited sample size of eyes with high myopia (SE ≤ -6 D; n = 22) in the previous literature. In addition, the duction angles differed between the two studies, with 20° of adduction in Liu et al.’s study compared with 30° of abduction in our study. Thus, although the absolute adduction-induced strain was not significantly higher than the abduction-induced strain in our myopic group, the strain per degree of ocular rotation may have been greater during adduction. This difference in duction angle should therefore be considered when directly comparing the biomechanical effects of adduction and abduction across studies. It is also crucial to note that myopic eyes with glaucoma exhibit distinct LC structural alterations, such as an increased prevalence of focal defects and large pores, compared to healthy myopic eyes, with these alterations potentially markedly altering the biomechanical responses of the LC ^[39]^ structures.

Interestingly, we observed no significant differences in global strains among the three mechanical loading conditions within the myopic group (P = 0.07), whereas significant differences were observed in the non-myopic group (P < 0.001). This finding suggests that myopic eyes may exhibit a more similar biomechanical response to different directions of ocular duction, potentially reflecting alterations in LC structure and material properties associated with myopia and glaucoma. At the tissue-special level, differences were primarily observed between acute IOP elevation and abduction within the PLT and LC, and these differences were far more pronounced in the non-myopic group. This blunted differential response in myopia might be attributed to an altered scleral biomechanics. Specifically, a more compliant sclera in highly myopic eyes may permit greater deformation of the peripapillary sclera and ONH boundary, thereby redistributing the applied load and potentially reducing local effective strain within the ONH during eye movements. Conversely, increased scleral stiffness may amplify these gaze-induced strains in emmetropic models.^[37, 40]^ Furthermore, axial elongation increases the tortuosity of the optic nerve in primary gaze position (due to the decreased availability of extraocular space in the posterior orbit).^[38]^ It may lead a relative indentation of parapapillary structures in primary gaze position (e.g., parapapillary temporal scleral ridges in highly myopic eyes). In adduction and abduction, however, the longer sagittal axis of the globe may lead to a more pronounced anterior movement of the oculat posterior pole in adduction and abduction, by which the optic nerve may become too short to allow the full movement, resulting in a backward pull by the optic nerve (dura mater) on the parapapillary structures, namely at the merging line of the dura mater with the posterior sclera.^[41, 42]^ Since the optic nerve exits the orbit in the nasal superior part of the orbit, the optic nerve backward pull may be the largest in the temporal inferior parapapillary region, which has the longest distance to the optic nerve exit out of the orbit.^[42]^ Fitting with this notion is the location of parapapillary suprachoroiudal cavitations, predominantly in the temporal inferior parapapillary region.^[42, 43]^ Taken together, increased scleral compliance and altered optic nerve and globe geometry may redistribute gaze-induced deformation over a broader region of the posterior eye, potentially contributing to the relatively similar global ONH strain magnitudes observed across loading conditions in the myopic group.

Additionally, we confirmed that severe baseline VF MD loss is a strong predictor of faster progression across both cohorts, which is consistent with the established understanding of glaucomatous disease dynamics.^[44]^

The limitations of our study should be acknowledged. First, patients in the myopic group were generally younger than those in the non-myopic group. Because age is a critical determinant of ocular biomechanical characteristics,^[45, 46]^ we rigorously evaluated its confounding potential. Based on our predefined statistical protocol, variables with a univariate P < 0.20 were included in multivariable analyses. Consequently, age was adjusted for in the myopic group’s multivariable model, ensuring our primary conclusions regarding ONH strains were controlled for age-related biomechanical variations. In the non-myopic group, age did not meet the statistical threshold for inclusion (univariate P = 0.36). Second, the cross-sectional nature of our OCT imaging limits our ability to track longitudinal biomechanical changes. Third, non-glaucomatous eyes as a control group were not included, neither for the non-myopic range nor for the myopic range. Finally, the majority of the myopic cohort had mild-to-moderate myopia (-0.5 to -6.0 diopters, 66.7%). While this limits direct generalizability to highly myopic populations, it highlights the need to explore whether a curvilinear association exists between axial length and biomechanical strain, particularly as structural degradation is known to accelerate beyond an axial length of 26.5 mm. Future research should utilize longitudinal imaging data and perform continuous regression analyses across a broader spectrum of axial lengths to validate and expand upon our observations.

In summary, our results indicate that an amplified strain response in the ONH during adduction serves as a critical biomechanical indicator of glaucoma progression in myopic NTG patients. Specifically, the exaggerated deformation observed within the PLT and choroid reflects the failure of load-bearing structures to buffer mechanical stress, resulting in the direct transmission of detrimental forces to these vulnerable, non-load-bearing neural and vascular tissues. This study highlights the profound influence of external, non-IOP-related mechanical forces on the physiology of the ONH and on the pathophysiology of myopic NTG and suggests to clinically incorporate ocular biomechanics into disease monitoring and management. Future large-scale, longitudinal studies are warranted to explore novel interventions aimed at alleviating biomechanical shear stress in myopic NTG eyes.

## Data Availability

All data produced in the present study are available upon reasonable request to the authors.

## CONFLICT OF INTEREST

No conflicting relationship exists for any author.

## FINANCLAL SUPPORT

This study was supported by the Program for Zhejiang Leading Talent of S&T Innovation (2021R52012), Key Research and Development Projects of Zhejiang Province (2022C03112), the National Natural Science Foundation of China (T2621006; 12472304; 12272030) and Beijing Natural Science Foundation (Z240017; L248028).

## Notes

### Competing Interest Statement

The authors have declared no competing interest.

### Author Declarations

This research complied with the ethical guidelines set forth in the Declaration of Helsinki and obtained authorization from the Ethics Committee of the Eye Hospital of Wenzhou Medical University (No. KYK-2013-12).

