## Supplementary material for "Association of Adduction-Induced Optic Nerve Head Strain with Visual Field Loss Progression in Myopic Normal-Tension Glaucoma *The Wenzhou Glaucoma Progression Study*": tables

| **Characteristic** | | **Overall**  **(N = 118)** | | **Myopic Group**  **(N = 66)** | | **Non-myopic Group**  **(N = 52)** | | **P value** |
| --- | --- | --- | --- | --- | --- | --- | --- | --- |
| **Participants** | | | | | | | | |
| Age at imaging, years | | 59.5 ±15.0 | | 54.0 ±15.3 | | 66.4 ±11.3 | | **< 0.001** |
| Gender, Male | | 51 (43.2) | | 36 (54.5) | | 15 (28.8) | | **0.009** |
| Follow-up, years | | 7.0 ±3.3 | | 6.5 ±3.4 | | 7.6±3.2 | | 0.09 |
| **Eyes** |  | |  | |  | |  | |
| Axial length, mm | | 24.5 ±1.7 | | 25.6 ±1.5 | | 23.2 ±0.6 | | **< 0.001** |
| Spherical equivalent, D | | -2.13 ±3.59 | | -4.58 ±2.93 | | 0.98 ±0.98 | | **< 0.001** |
| CCT, μm | | 531.0 ±33.4 | | 530.7 ±32.9 | | 531.5 ±34.4 | | 0.54 |
| IOP at imaging, mmHg | | 14.6 ±2.2 | | 14.5 ±2.4 | | 14.6 ±2.0 | | 0.78 |
| MD at imaging, dB | | -8.47 ±6.15 | | -8.99 ±6.21 | | -7.81 ±6.09 | | 0.33 |
| DH, N (%) | | 26 (22.0) | | 9 (13.6) | | 17 (32.7) | | 0.17 |

**Table 1. Demographics and Ocular Characteristics in Normal Tension Glaucoma Patients between Myopic and Non-Myopic Group.** CCT = central cornea thickness, IOP = intraocular pressure, MD = mean deviation, DH = disc hemorrhage. Significant P values are shown in bold type.

| **Characteristic** | **Strains induced by IOP** | **Strains induced by AD** | **Strains induced by AB** | **P value** | | **Post Hoc Significance** |
| --- | --- | --- | --- | --- | --- | --- |
| Myopic group, (%) | 10.1 ±2.7 | 9.8 ±2.7 | 9.2 ±2.3 | | 0.07 | 0.76,  0.06, 0.26 |
| Non-myopic group, (%) | 9.3 ±1.8 | 9.9 ±2.0 | 8.4 ±2.1 | | **<0.001** | 0.20,  **0.026**, **< 0.001** |

**Table 2. Effective strains in Myopic and Non-myopic Group.** IOP = intraocular pressure, AD = adduction, AB = abduction. Significant P values are shown in bold type. Post hoc significant differences are shown as “>”.

Post Hoc Significance: IOP-AD, IOP -AB, AD- AB

| **Characteristic** | **Strains induced by IOP** | **Strains induced by AD** | **Strains induced by AB** | **P value** | **Post Hoc Significance** |
| --- | --- | --- | --- | --- | --- |
| **Myopic group** |  |  |  |  |  |
| Sclera, (%) | 9.4 ±2.7 | 9.0 ±2.6 | 8.7 ±2.6 | 0.23 | 0.59, 0.21, 0.75 |
| Choroid, (%) | 9.7 ±3.1 | 9.3 ± 2.7 | 8.8 ±2.6 | 0.10 | 0.65, 0.08, 0.42 |
| Pre-Lamina Tissue, (%) | 10.6 ±2.7 | 10.5 ±2.8 | 9.6 ±2.3 | **0.036** | 0.90, **0.044**, 0.12 |
| Lamina Cribrosa, (%) | 11.3 ±2.9 | 11.3 ±3.1 | 10.3 ±2.8 | **0.015** | 1.00, **0.035**, **0.036** |
| **Non-myopic group** |  |  |  |  |  |
| Sclera, (%) | 8.6 ±2.0 | 8.9 ±2.0 | 7.8 ±2.2 | **0.005** | 0.57, 0.08, **0.005** |
| Choroid, (%) | 8.3 ±1.7 | 8.9 ±2.0 | 7.4 ±2.0 | **< 0.001** | 0.10, **0.023**, **< 0.001** |
| Pre-Lamina Tissue, (%) | 9.8 ±1.9 | 10.5 ±2.2 | 8.9 ±2.3 | **0.001** | 0.17, **0.040**, **0.001** |
| Lamina Cribrosa, (%) | 10.4 ±2.2 | 11.3 ±2.5 | 9.1 ±2.3 | **< 0.001** | 0.10, **0.010,** **< 0.001** |

**Table 3. Effective Strains under Various Conditions in Myopic and Non-myopic Group.** IOP = intraocular pressure, AD = adduction, AB = abduction. Significant P values are shown in bold type. Post hoc significant differences are shown as “>”.

Post Hoc Significance: IOP-AD, IOP -AB, AD- AB

|  | **Myopic Group** | | | | **Non-myopic Group** | | | | |
| --- | --- | --- | --- | --- | --- | --- | --- | --- | --- |
|  | **Univariate Model** | | **Multivariable Model** | | **Univariate Model** | | | **Multivariable Model** | |
| **Characteristic** | **Coefficient (dB/yr)** | **P value** | **Coefficient (dB/yr)** | **P value** | **Coefficient (dB/yr)** | **P value** | **Coefficient (dB/yr)** | | **P value** |
| Age at imaging, years | 0.032 | **0.046** | 0.009 | 0.58 | 0.023 | 0.36 |  | |  |
| Sex, Male | -0.285 | 0.60 |  |  | 0.134 | 0.82 |  | |  |
| Axial length, mm | -0.288 | **0.09** |  |  | 0.155 | 0.72 |  | |  |
| Spherical equivalent, D | 0.175 | **0.043** | 0.085 | 0.33 | 0.130 | 0.61 |  | |  |
| CCT, μm | -0.001 | 0.90 |  |  | 0.005 | 0.54 |  | |  |
| IOP at imaging, mmHg | 0.039 | 0.72 |  |  | -0.169 | 0.19 |  | |  |
| MD at imaging, dB | 0.470 | **< 0.001** | 0.397 | **< 0.001** | 0.263 | **< 0.001** | 0.249 | | **< 0.001** |
| DH, N (%) | -0.188 | 0.75 |  |  | 0.154 | 0.75 |  | |  |
| Strains induced by IOP, (%) | 0.179 | **0.017** | 0.184 | **0.004** | 0.134 | 0.24 |  | |  |
| Strains induced by adduction, (%) | -0.142 | **0.07** | -0.139 | **0.025** | 0.110 | 0.26 |  | |  |
| Strains induced by abduction, (%) | 0.099 | 0.28 |  |  | 0.156 | 0.11 | 0.076 | | 0.39 |

**Table 4. Linear Mixed-Effects Regression Models Evaluating the Effect of Characteristics and Effective Strains under Various Conditions in the Rates of Mean Deviation Loss Over Time.** CCT = central cornea thickness, IOP = intraocular pressure, MD = mean deviation, DH = disc hemorrhage.

|  | **Multivariable Model 1** | | **Multivariable Model 2** | | **Multivariable Model 3** | | **Multivariable Model 4** | |
| --- | --- | --- | --- | --- | --- | --- | --- | --- |
| **Characteristic** | **Coefficient (dB/yr)** | **P value** | **Coefficient (dB/yr)** | **P value** | **Coefficient (dB/yr)** | **P value** | **Coefficient (dB/yr)** | **P value** |
| IOP |  |  |  |  |  |  |  |  |
| Age at imaging, years | 0.005 | 0.76 | 0.009 | 0.59 | 0.009 | 0.57 | 0.010 | 0.57 |
| Spherical equivalent, D | 0.087 | 0.32 | 0.075 | 0.41 | 0.068 | 0.46 | 0.054 | 0.55 |
| MD at imaging, dB | 0.418 | **< 0.001** | 0.461 | **< 0.001** | 0.445 | **< 0.001** | 0.501 | **< 0.001** |
| Sclera, (%) | 0.203 | 0.002 |  |  |  |  |  |  |
| Choroid, (%) |  |  | 0.174 | **0.002** |  |  |  |  |
| Pre-Lamina Tissue, (%) |  |  |  |  | 0.135 | **0.021** |  |  |
| Lamina Cribrosa, (%) |  |  |  |  |  |  | 0.147 | **0.015** |
| AD |  |  |  |  |  |  |  |  |
| Age at imaging, years | 0.015 | 0.38 | 0.013 | 0.42 | 0.014 | 0.40 | 0.013 | 0.45 |
| Spherical equivalent, D | 0.025 | 0.78 | 0.023 | 0.79 | 0.028 | 0.75 | 0.029 | 0.75 |
| MD at imaging, dB | 0.410 | **< 0.001** | 0.411 | **< 0.001** | 0.407 | **< 0.001** | 0.410 | **< 0.001** |
| Sclera, (%) | -0.111 | 0.10 |  |  |  |  |  |  |
| Choroid, (%) |  |  | -0.144 | **0.031** |  |  |  |  |
| Pre-Lamina Tissue, (%) |  |  |  |  | -0.135 | **0.033** |  |  |
| Lamina Cribrosa, (%) |  |  |  |  |  |  | -0.070 | 0.17 |

**Table 5. Linear Mixed-Effects Regression Models Evaluating the Effect of Characteristics and Tissue-specific Effective Strains under Various Conditions in the Rates of Mean Deviation Loss Over Time** **in the myopic group.** CCT = central cornea thickness, IOP = intraocular pressure, MD = mean deviation, DH = disc hemorrhage.
