## Supplementary Materials for "Association of Adduction-Induced Optic Nerve Head Strain with Visual Field Loss Progression in Myopic Normal-Tension Glaucoma *The Wenzhou Glaucoma Progression Study*"

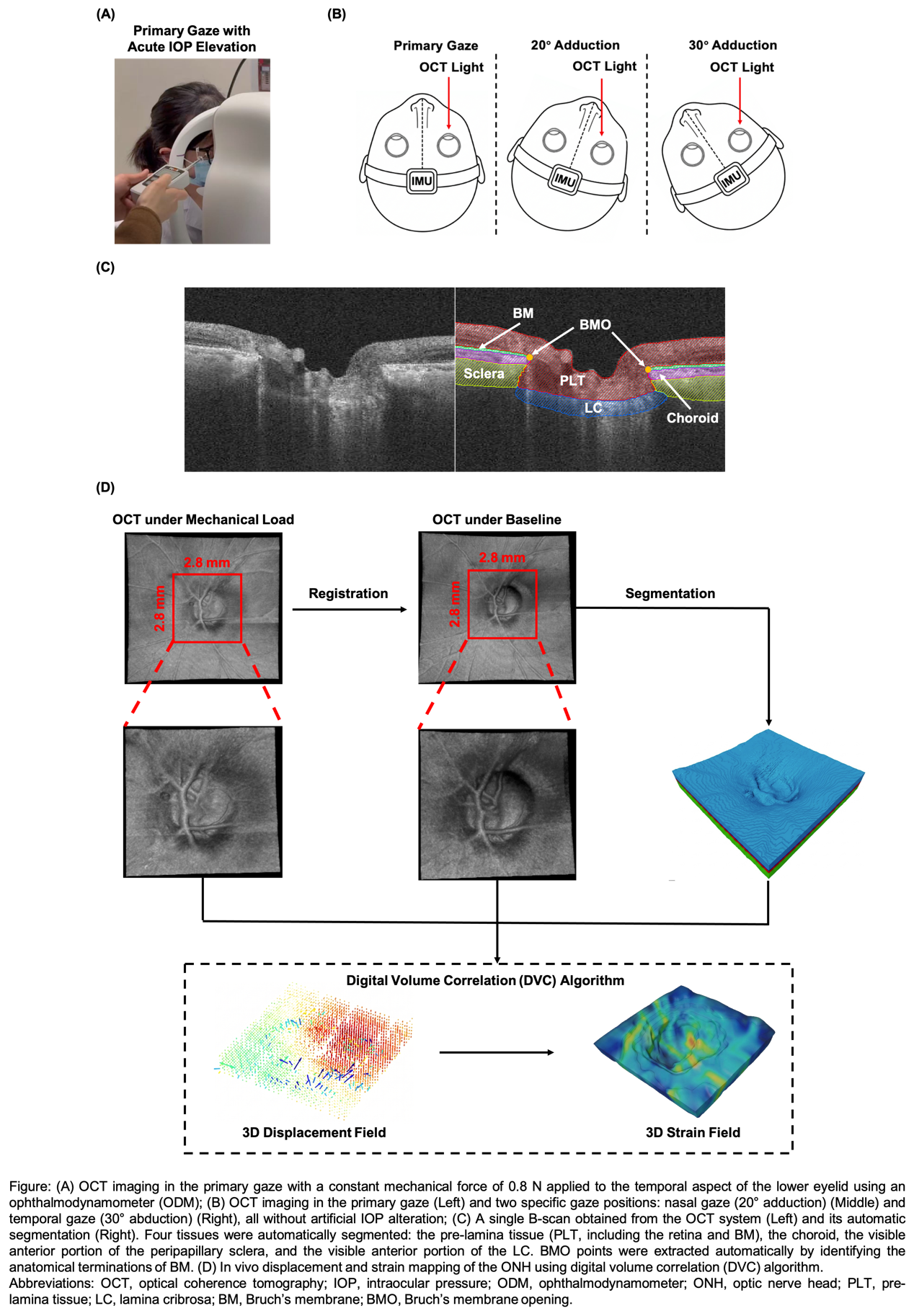

**Supplementary Materials**

**Supplementary Details for Automatic Segmentation……………………..……….1**

**Supplementary Details for Three-Dimensional Strain Mapping………………....3**

**Supplementary Details for Validation of DVC Algorithm………………………..4**

**Reference…………………………………………………………………………….11**

**Supplementary Details for Automatic Segmentation**

Automatic Optic Nerve Head (ONH) Segmentation

Segmentation of ONH tissues was performed by fine-tuning SAM-Med2D,^1^ a medical-image adaptation of the Segment Anything Model (SAM). SAM-Med2D was developed to reduce the domain gap between natural-image segmentation and medical-image segmentation by fine-tuning SAM on a large-scale two-dimensional medical image dataset with multiple prompt types. The model was selected because its pretrained image encoder provides a generalizable feature representation, while task-specific fine-tuning allows adaptation to ONH OCT images acquired in this study.

Fine-Tuning Dataset

The fine-tuning dataset was derived from ONH OCT images acquired using the same swept-source OCT system as in the main study (VG200D; SVision Imaging Ltd., Henan, China). The system uses a central wavelength of approximately 1050 nm and operates at 200,000 A-scans per second. Each volumetric scan covered a 6 × 6 mm region centered on the ONH and consisted of 512 serial B-scans with 512 A-scans per B-scan. The mean spacing between adjacent B-scans was 11.7 μm, with average axial and lateral pixel resolutions of 3.23 μm and 11.7 μm, respectively.

To improve the diversity of the training data, 100 B-scan slices were randomly selected from each three-dimensional OCT volume. Manual annotations were performed for five anatomical labels: pre-lamina tissue (PLT, including the retina), Bruch’s membrane (BM), choroid, the visible anterior portion of the peripapillary sclera, and the visible anterior portion of the lamina cribrosa (LC). The dataset contained 100 eyes and 10,000 manually annotated B-scans in total. The images were divided into training (fine-tuning), validation, and testing sets at a 3:1:1 ratio.

Fine-Tuning Procedure

Fine-tuning followed empirical recommendations for adapting foundation models to medical image segmentation.^2^ Briefly, the Vision Transformer-Large (ViT-L) backbone was used, and parameter-efficient fine-tuning (PEFT) was applied to both the image encoder and the mask decoder. The validation set was used during model development, and segmentation performance was evaluated on the held-out testing set. The trained model was then applied slice-by-slice to ONH OCT volumes, and the two-dimensional segmentations were stacked to reconstruct three-dimensional tissue volumes.

The resulting automatic segmentations were used in two ways. First, the PLT, choroid, visible anterior peripapillary sclera, and visible anterior LC defined tissue domains for tissue-specific strain aggregation. Second, BM was segmented independently to identify Bruch’s membrane opening (BMO) points; the BMO points defined the BMO plane, which served as the standardized horizontal reference plane for each ONH.

Segmentation Performance

Segmentation accuracy was evaluated using the Dice coefficient and intersection-over-union (IoU),^3,4^ with performance summarized in **Table 1**. The model achieved high segmentation performance for PLT, choroid, and sclera, with lower but acceptable performance for BM and LC, which are thinner or less consistently visible in OCT images.

*Table 1. Segmentation performance of the fine-tuned SAM-Med2D model.*

|  | PLT | BM | Choroid | Sclera | LC |
| --- | --- | --- | --- | --- | --- |
| Dice | 0.973 | 0.839 | 0.933 | 0.922 | 0.784 |
| Iou | 0.948 | 0.725 | 0.876 | 0.859 | 0.666 |

Abbreviations: PLT = pre-lamina tissue; BM = Bruch’s membrane; LC = lamina cribrosa; IoU = intersection-over-union.

**Supplementary Details for Three-Dimensional Strain Mapping**

Details of DVC Algorithm

The OCT-based DVC workflow used a two-stage local-to-global approach. Therefore, only a brief overview is provided here. For each eye, the baseline primary-gaze OCT volume was paired with the corresponding loaded volume acquired during acute IOP elevation, 20° adduction, or 30° abduction. After adaptive compensation and rigid registration, DVC was performed within the BMO-centered analysis region to estimate three-dimensional tissue displacement.

In the first stage, a subset-based local DVC analysis was performed using the DVC Local Approach module in the XDigitalVolumeCorrelation Extension of Amira. An isotropic sub-volume size of 30 μm was used. Image similarity between corresponding sub-volumes was assessed using the correlation metric, with a correlation threshold of 0.7, and the transformation model permitted translation and rotation. This local analysis was used to capture the initial displacement field, particularly in regions exhibiting relatively large motion.

The displacement field derived from the local DVC analysis was subsequently used as the initial displacement estimate for the finite-element-based global DVC analysis. In the global approach, the region of interest was represented by a hexahedral finite-element mesh, and the displacement field was iteratively optimized by minimizing the gray-scale intensity difference between the reference and deformed OCT volumes. For each OCT volume, an isotropic mesh size of 100 × 100 × 100 μm was used, corresponding to approximately 9 × 31 × 9 voxels in the X, Y, and Z directions, respectively. The convergence tolerance of δu was set to 0.01 μm. The resulting global nodal displacement field was used for subsequent strain derivation and tissue-specific aggregation.

Derivation of Effective Strain

The spatial displacement gradient was calculated from the DVC-derived displacement field. The deformation gradient was defined as F = I + ∇u, where I is the identity tensor and ∇u is the displacement gradient. The Green-Lagrange strain tensor was then calculated as E = 1/2(FᵀF − I). The three principal components of E were denoted as E₁, E₂, and E₃.

Effective strain was used as the main scalar summary of local three-dimensional deformation. This measure is a single positive index that reflects both tensile and compressive deformation, allowing complex ONH tissue deformation to be summarized independent of direction. Effective strain ($E_{eff}$) was calculated from the principal Green-Lagrange strain components as:

$$E_{eff} =\sqrt{\frac{{(E_{1}-E_{2})}^{2}+{(E_{1}-E_{3})}^{2}+{(E_{2}-E_{3})}^{2}}{2}}$$

For global ONH analysis, local effective strains were averaged within the cropped ONH analysis region. For tissue-specific analysis, local effective strains were averaged separately within the automatically segmented PLT, choroid, sclera, and LC volumes. BM was not included as a strain-analysis tissue; it was used to define BMO and the BMO reference plane.

**Supplementary Details for Validation of DVC Algorithm**

Synthetic Deformation Test

To validate the accuracy of the DVC algorithm, four baseline primary-gaze OCT volumes were randomly selected and deformed using a known synthetic three-dimensional bell-shaped displacement field. The DVC workflow used in the main analysis was then applied to each baseline-deformed volume pair. The DVC-derived displacement and effective strain fields were compared with the known ground-truth fields generated from the imposed deformation.

The imposed displacement was applied along the Y axis, corresponding to the axial image dimension. The peak displacement was set to 100 μm at the center of the field. A Gaussian width parameter of σ = 70 μm was used so that the displacement gradually decayed toward the boundary of the region of interest, reaching approximately 10 μm near the edge. The displacement field was defined as:

$u_{y}\left( x,y,z \right)=A\cdot\exp\left( -\frac{\left( x-x_{0} \right)^{2}+\left( y-y_{0} \right)^{2}+\left( z-z_{0} \right)^{2}}{2\sigma^{2}} \right)$,

$u_{x}\left( x,y,z \right)=0$,

$u_{z}\left( x,y,z \right)=0$.

where A = 100 μm, σ = 70 μm, and ($x_{0}$, $y_{0}$, $z_{0}$) denotes the center of the synthetic displacement field.

Displacement accuracy was assessed by calculating the mean DVC-derived displacement in the X, Y, and Z directions and the corresponding mean absolute error (MAE) against the ground truth, with results summarized in **Table 2**. Because the imposed deformation had a focal high-displacement region, we also evaluated the mean axial displacement and MAE within the top 20% of voxels ranked by ground-truth Y-axis displacement. Spatial agreement of the high-displacement region was quantified using the Dice coefficient after binarizing the top-20% region as 1 and all remaining voxels as 0.

Across the four validation cases, the off-axis displacements remained close to zero, with overall mean X- and Z-axis displacements of -0.01 ± 1.22 μm and 0.05 ± 1.55 μm, respectively. The corresponding overall MAEs were 0.26 ± 1.06 μm for X displacement and 0.24 ± 1.29 μm for Z displacement, indicating minimal displacement leakage into non-imposed directions. For the imposed Y-axis displacement, the overall MAE was 2.48 ± 2.49 μm. In the top-20% high-displacement region, the mean DVC-derived Y-axis displacement was 66.88 ± 11.01 μm, the MAE was 3.89 ± 3.72 μm, and the Dice coefficient was 0.92, demonstrating strong agreement between the DVC-derived and ground-truth high-displacement regions.

Effective strain accuracy was evaluated using the same ground-truth deformation field, with results summarized in **Table 3**. For each case, we summarized the mean, median, and 95th-percentile effective strain, as well as the MAE of effective strain across the full region of interest. The top-20% high-strain region was further assessed using mean effective strain, MAE, and Dice overlap with the ground-truth high-strain region.

The DVC-derived effective strain closely reproduced the imposed deformation pattern. Across all cases, the mean effective strain was 4.60 ± 2.68%, the median was 4.43%, and the 95th percentile was 7.83%. The overall effective-strain MAE was 0.70 ± 1.51%. Within the top-20% high-strain region, the mean effective strain was 8.16 ± 2.28%, the MAE was 1.38 ± 2.93%, and the Dice coefficient was 0.76. Although high-strain-region overlap was lower than high-displacement-region overlap, this is expected because strain is derived from spatial gradients of displacement and is therefore more sensitive to local interpolation and numerical differentiation.

*Table 2. Accuracy of DVC-derived displacement under the synthetic bell-shaped displacement field. Values are mean ± SD in μm except for Dice coefficients.*

|  | Disp_X | MAE_Disp_X | Disp_Z | MAE_Disp_Z | Disp_Y | MAE_Disp_Y | Disp_Y_20% | MAE_Disp_Y_20% | Dice_Disp_Y_20% |
| --- | --- | --- | --- | --- | --- | --- | --- | --- | --- |
| Case 1 | 0.02±2.41 | 0.26±2.29 | 0.02±3.07 | 0.25±2.96 | 29.11±24.20 | 2.45±2.00 | 70.97±11.38 | 3.51±2.47 | 0.93 |
| Case 2 | 0.01±0.57 | 0.20±0.45 | 0.02±0.69 | 0.18±0.32 | 26.09±20.95 | 2.23±1.75 | 63.51±11.16 | 3.05±2.09 | 0.92 |
| Case 3 | 0.02±0.69 | 0.22±0.50 | 0.04±1.27 | 0.22±0.83 | 27.21±22.92 | 2.37±1.90 | 67.77±11.29 | 3.43±2.43 | 0.92 |
| Case 4 | -0.09±1.21 | 0.34±1.00 | 0.18±1.17 | 0.32±1.03 | 26.94±22.10 | 2.85±4.30 | 65.29±10.21 | 5.57±7.91 | 0.89 |
| Overall (Mean) | -0.01±1.22 | 0.26±1.06 | 0.05±1.55 | 0.24±1.29 | 27.34±22.54 | 2.48±2.49 | 66.88±11.01 | 3.89±3.72 | 0.92 |

Abbreviations: Disp_X, Disp_Y, and Disp_Z = DVC-derived displacement along the X, Y, and Z axes; MAE_Disp_X, MAE_Disp_Y, and MAE_Disp_Z = mean absolute error for each displacement component; Disp_Y_20% and MAE_Disp_Y_20% = mean displacement and MAE within the top-20% high-Y-displacement region; Dice_Disp_Y_20% = Dice overlap of that high-displacement region.

*Table 3. Accuracy of DVC-derived effective strain under the synthetic bell-shaped displacement field. Values are mean ± SD in %,* *except for medians, 95th percentiles, and Dice coefficients.*

|  | Mean_ES | Median_ES | 95%th_ES | MAE_ES | ES_20% | MAE_ES_20% | Dice_ES_20% |
| --- | --- | --- | --- | --- | --- | --- | --- |
| Case 1 | 4.65±2.57 | 4.54 | 7.92 | 0.61±1.24 | 8.05±1.92 | 0.97±2.42 | 0.78 |
| Case 2 | 4.45±2.41 | 4.44 | 7.42 | 0.61±1.20 | 7.61±1.63 | 1.12±2.45 | 0.77 |
| Case 3 | 4.50±2.52 | 4.38 | 7.68 | 0.64±1.35 | 7.88±1.85 | 1.21±2.80 | 0.77 |
| Case 4 | 4.79±3.22 | 4.40 | 8.29 | 0.94±2.26 | 9.11±3.70 | 2.21±4.06 | 0.72 |
| Overall (Mean) | 4.60±2.68 | 4.43 | 7.83 | 0.70±1.51 | 8.16±2.28 | 1.38±2.93 | 0.76 |

Abbreviations: ES = effective strain; Mean_ES, Median_ES, and 95%th_ES = mean, median, and 95th-percentile effective strain; MAE_ES = mean absolute error of effective strain; ES_20% and MAE_ES_20% = mean effective strain and MAE within the top-20% high-strain region; Dice_ES_20% = Dice overlap of that high-strain region.

Parameter Sensitivity Analysis

We further examined whether the selected DVC parameters provided a reasonable balance between accuracy and computational stability. The parameter set used in the main study was a 100 × 100 × 100 μm hexahedral mesh, a convergence tolerance of δu = 0.01 μm, and a 256-slice cropped volume. This setting was compared with three alternatives: a stricter convergence tolerance (100 μm mesh, δu = 0.001 μm, 256 slices), a finer mesh (50 μm mesh, δu = 0.01 μm, 256 slices), and a reduced number of slices (100 μm mesh, δu = 0.01 μm, 128 slices). In Figures 1–3 and 5–7, the x-axis categories denote the prescribed ground truth (GT) and DVC parameter settings in the order of mesh size, convergence tolerance, and number of slices used for the cropped volume.

For displacement estimation, the selected 100 μm mesh with δu = 0.01 μm and 256 slices closely matched the ground truth across the global mean Y-axis displacement, top-20% Y-axis displacement, and high-displacement-region Dice overlap (**Figures 1–3**). It reproduced the global mean Y-axis displacement (27.34 ± 1.10 μm vs 29.46 ± 0.96 μm for ground truth), the top-20% Y-axis displacement (66.88 ± 2.81 μm vs 67.47 ± 2.61 μm), and high-displacement-region overlap (Dice = 0.92 ± 0.02). Tightening the convergence tolerance to 0.001 μm produced similar results. In contrast, the 50 μm mesh underestimated displacement and reduced Dice overlap, while the 128-slice cropped volume showed slightly lower top-20% displacement and Dice overlap. **Figure 4** further illustrates this agreement spatially: the red top-20% high-displacement regions derived from DVC largely colocalized with the ground-truth high-displacement regions, whereas most of the remaining tissue was correctly classified as non-high-displacement tissue. These results support the selected displacement-tracking parameters.

For effective strain, the selected parameter set also provided stable and accurate estimates across global effective-strain summaries, top-20% high-strain values, and high-strain-region Dice overlap (**Figures 5–7**). It reproduced the ground-truth mean, median, and 95th-percentile effective strain (mean: 4.60 ± 0.13% vs 4.81 ± 0.06%; median: 4.43 ± 0.06% vs 4.71 ± 0.07%; 95th percentile: 7.83 ± 0.32% vs 7.75 ± 0.08%). The top-20% effective strain was similarly close to the ground truth (8.16 ± 0.66% vs 8.14 ± 0.29%). The stricter convergence tolerance again produced comparable results. The 50 μm mesh generated inflated 95th-percentile and top-20% effective strain values, and both the 50 μm mesh and 128-slice setting reduced Dice overlap for high-strain regions. **Figure 8** shows representative high-strain maps; although the top-20% strain overlap was lower than the displacement overlap, the DVC-derived high-strain regions captured the principal spatial pattern of the ground-truth high-strain region. These findings indicate that the selected parameters minimize numerical instability while preserving the magnitude and spatial distribution of the imposed deformation.

Together, these validation experiments demonstrate that the DVC workflow used in this study accurately recovers both the magnitude and spatial distribution of imposed three-dimensional ONH deformation. The low displacement and effective-strain errors, high Dice overlap for high-displacement regions, and parameter-sensitivity results support the reliability of the selected DVC settings for quantifying ONH strains in the main analysis.

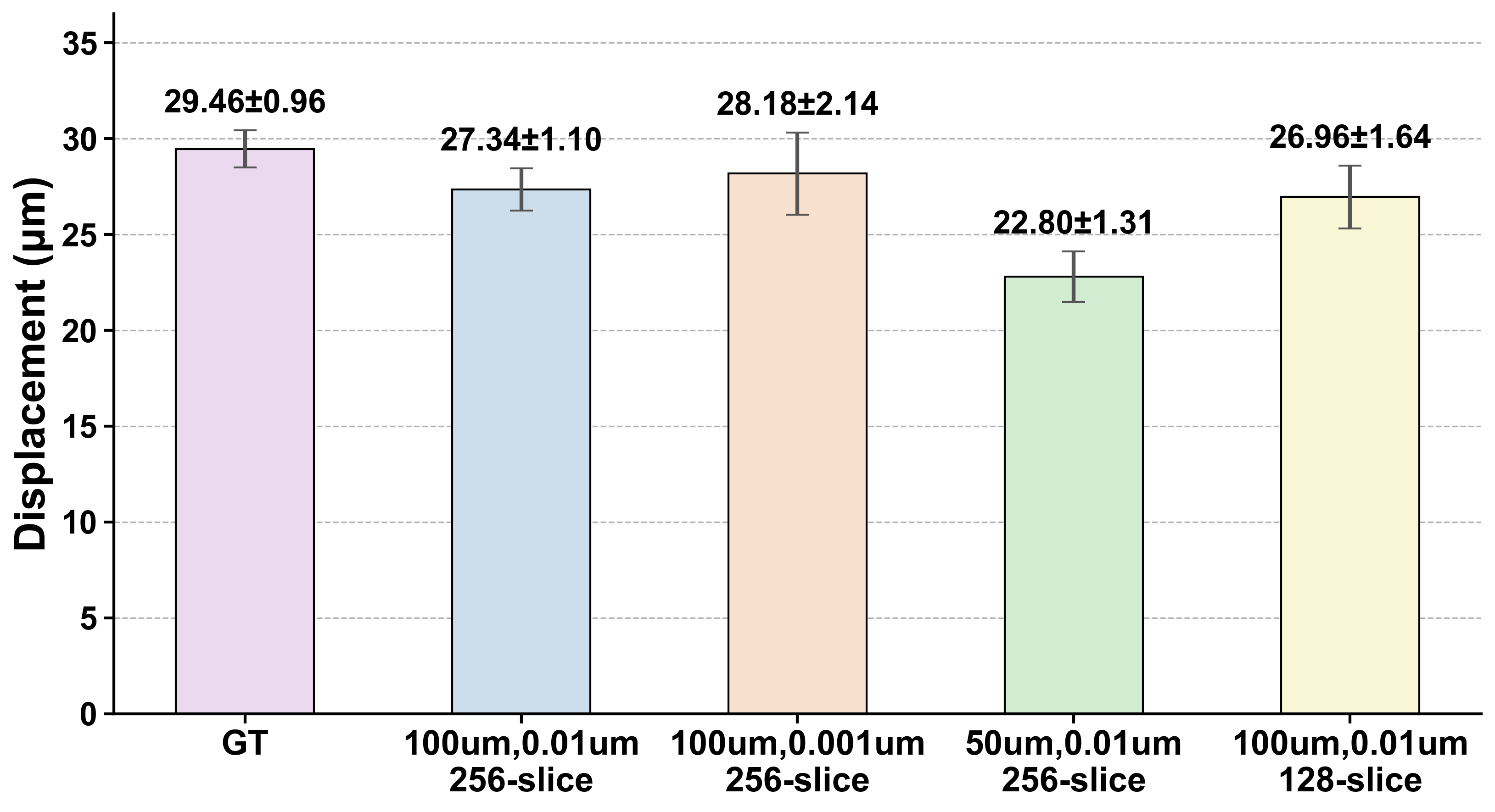

Figure 1. Global mean Y-axis displacement across DVC parameter settings. Bars show mean ± SD across the four validation cases. The x-axis shows GT and DVC settings formatted as mesh size, convergence tolerance, and cropped-volume slice number. GT = ground truth.

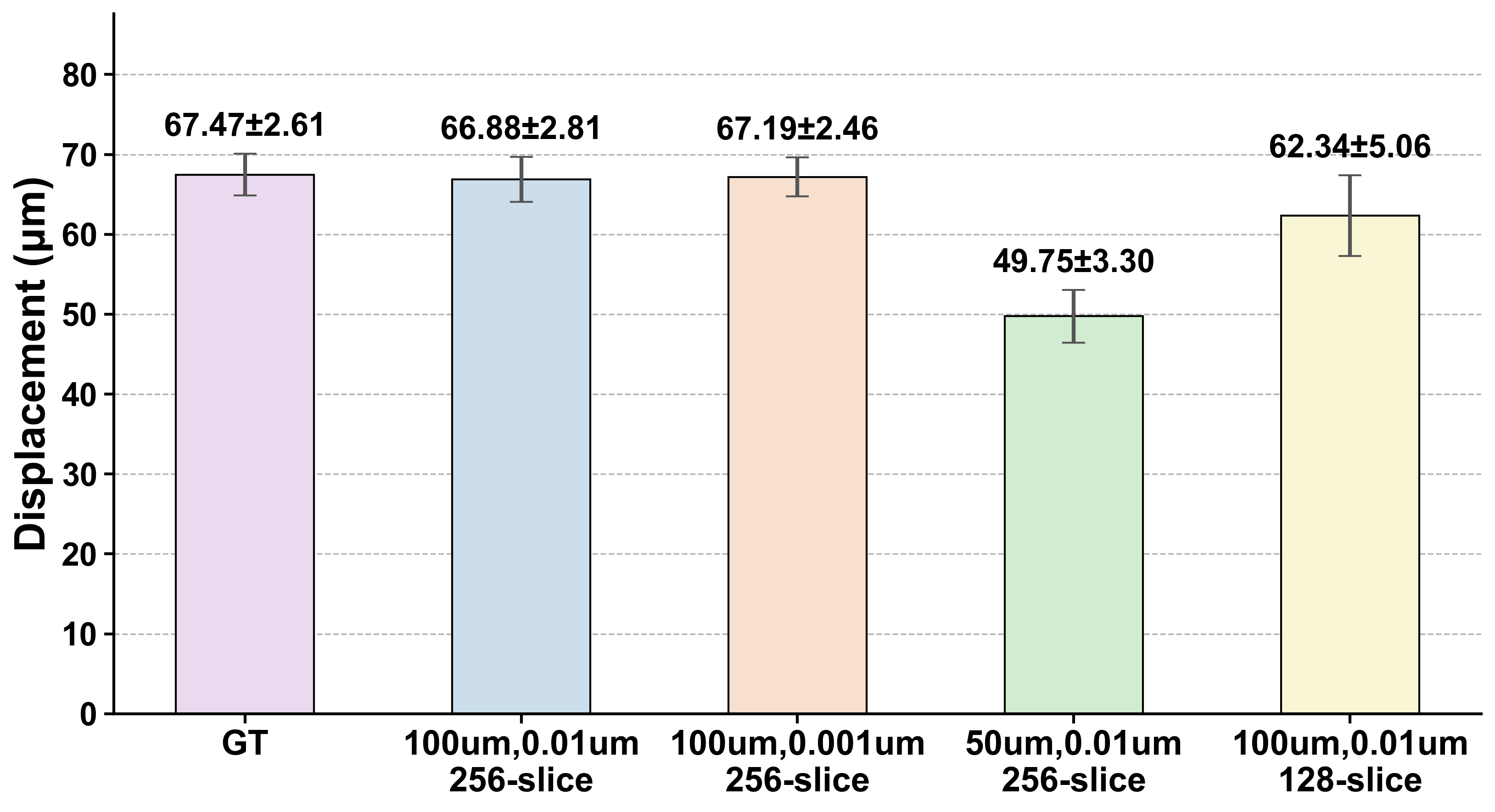

Figure 2. Mean Y-axis displacement within the top-20% high-displacement region across DVC parameter settings. Bars show mean ± SD across the four validation cases. The x-axis shows GT and DVC settings formatted as mesh size, convergence tolerance, and cropped-volume slice number. GT = ground truth.

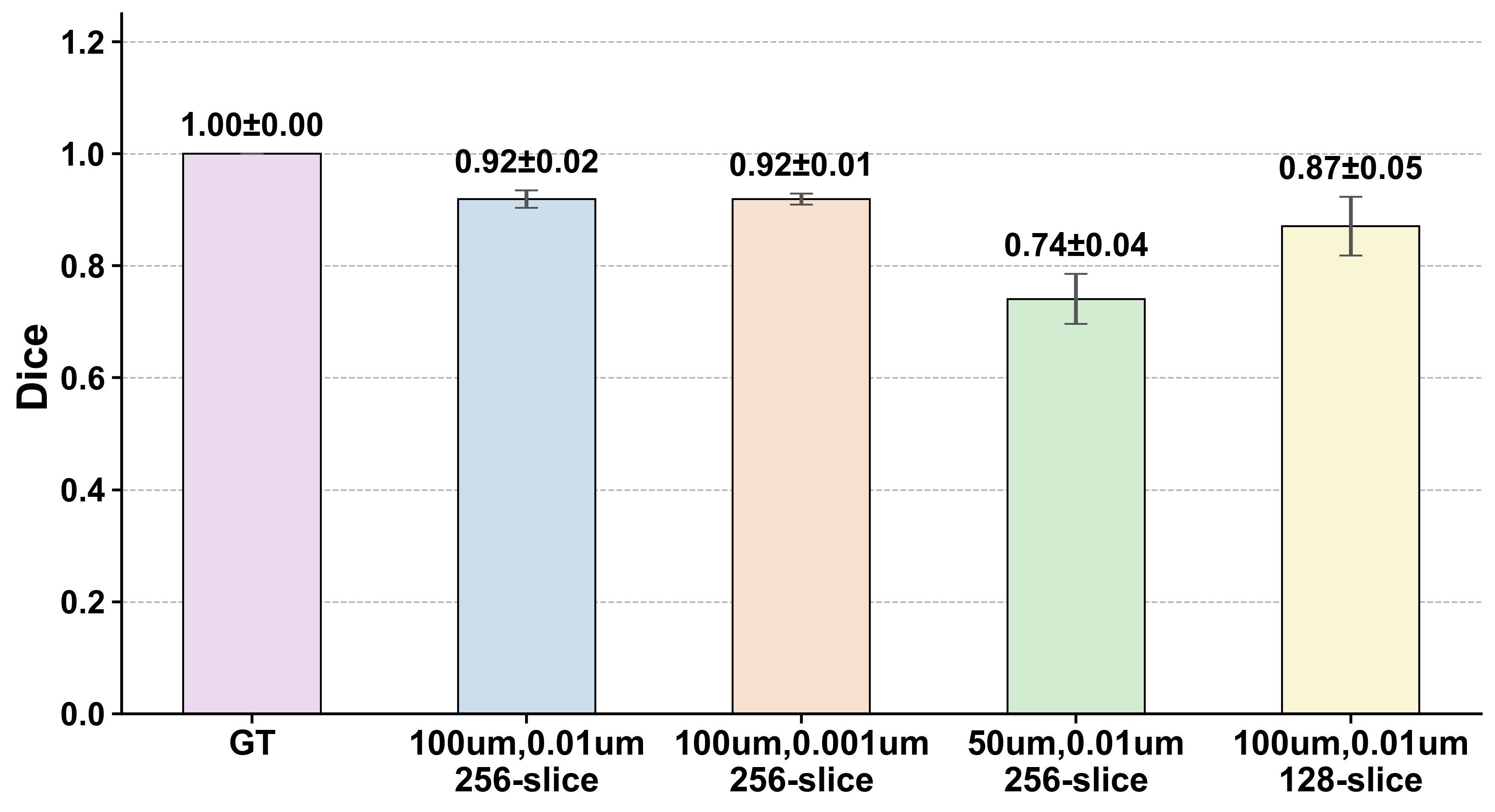

Figure 3. Dice overlap of the top-20% high-displacement region between the ground truth and each DVC parameter setting. Bars show mean ± SD across the four validation cases. The x-axis shows GT and DVC settings formatted as mesh size, convergence tolerance, and cropped-volume slice number.

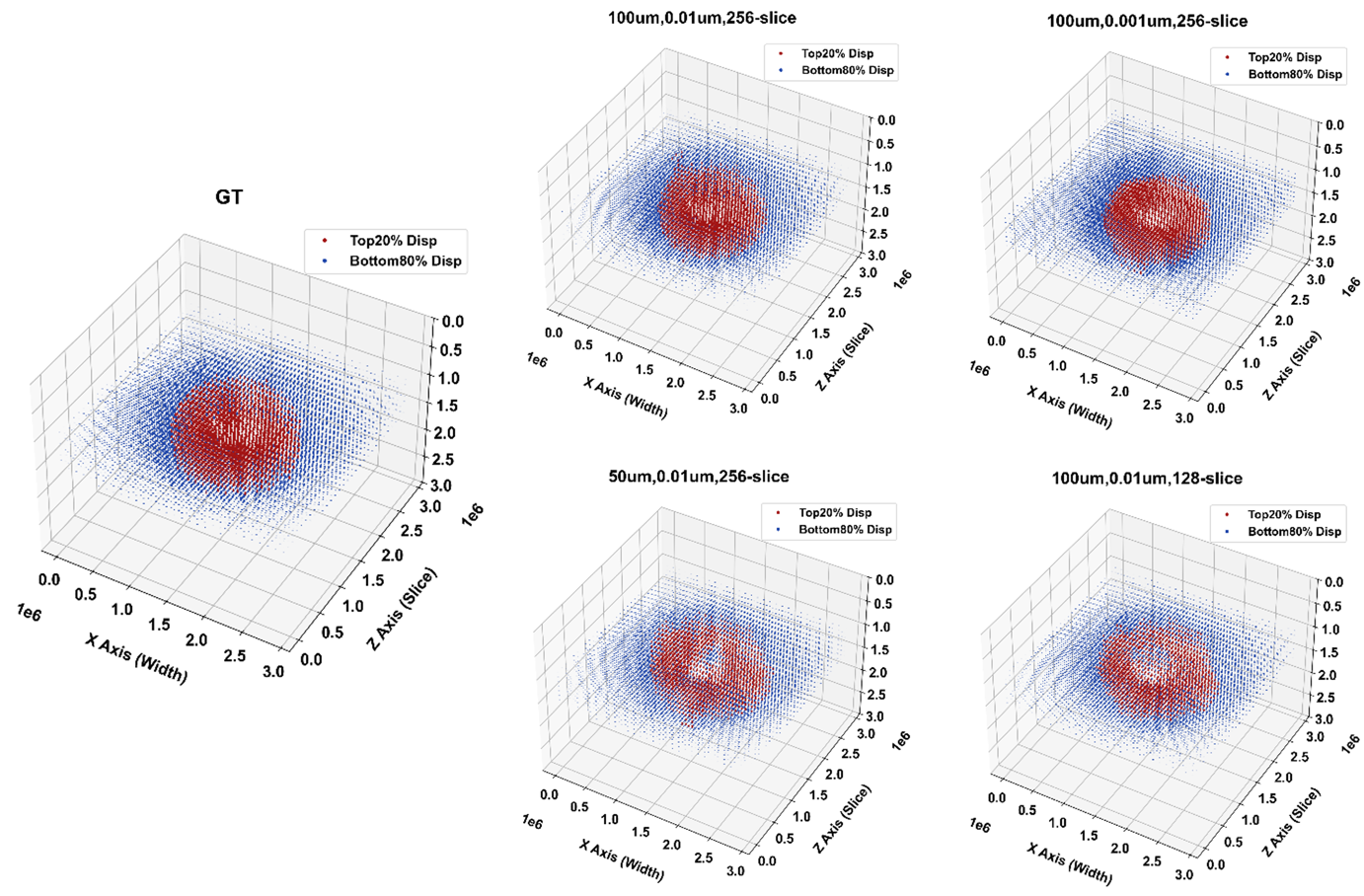

Figure 4. Representative spatial maps of top-20% high-displacement regions. Red indicates the high-displacement region and blue indicates the remaining region, allowing visual comparison between the ground truth and DVC-derived displacement maps.

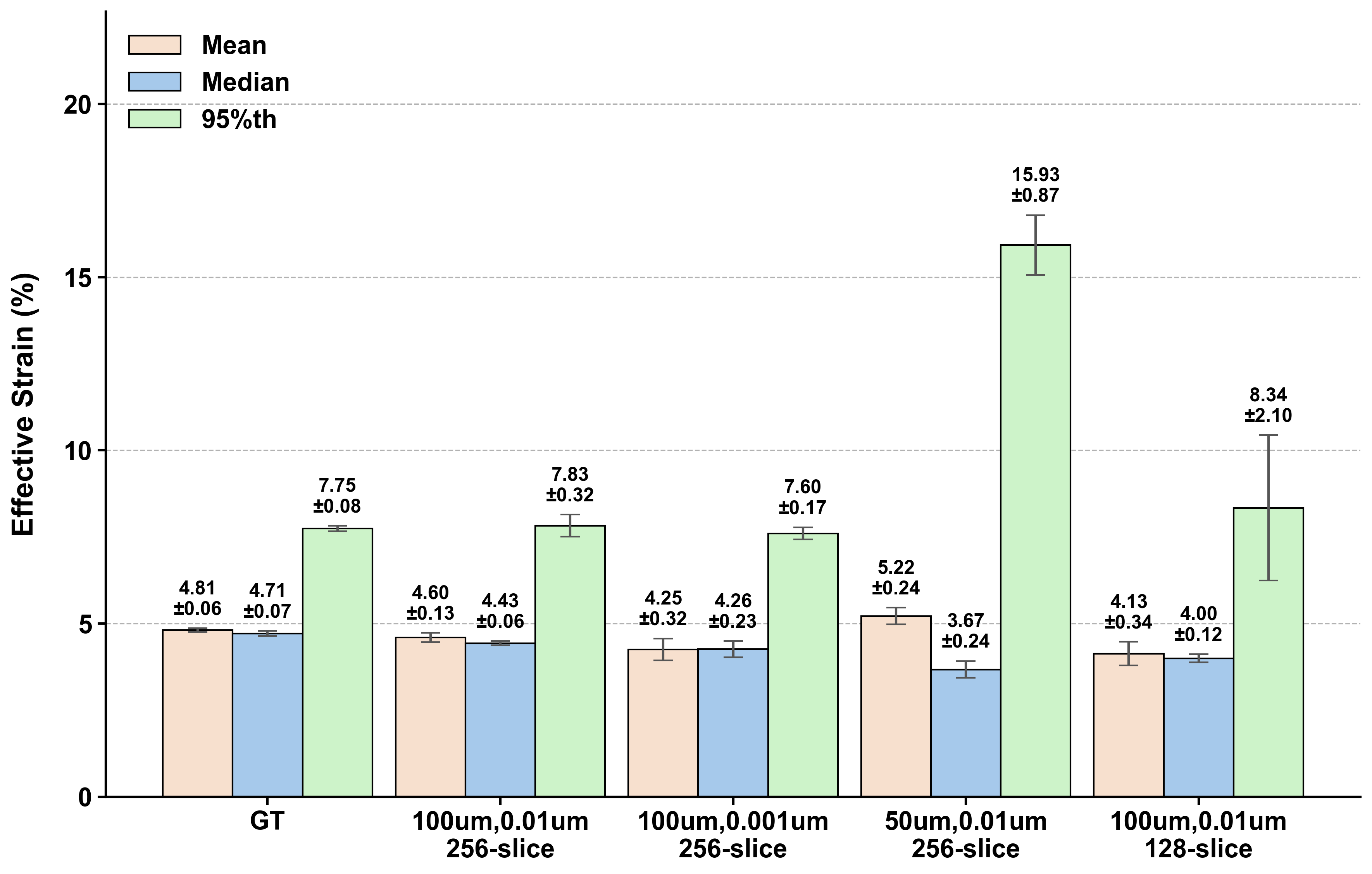

Figure 5. Global effective-strain summary across DVC parameter settings, including mean, median, and 95th-percentile effective strain. Bars show mean ± SD across the four validation cases. The x-axis shows GT and DVC settings formatted as mesh size, convergence tolerance, and cropped-volume slice number. GT = ground truth.

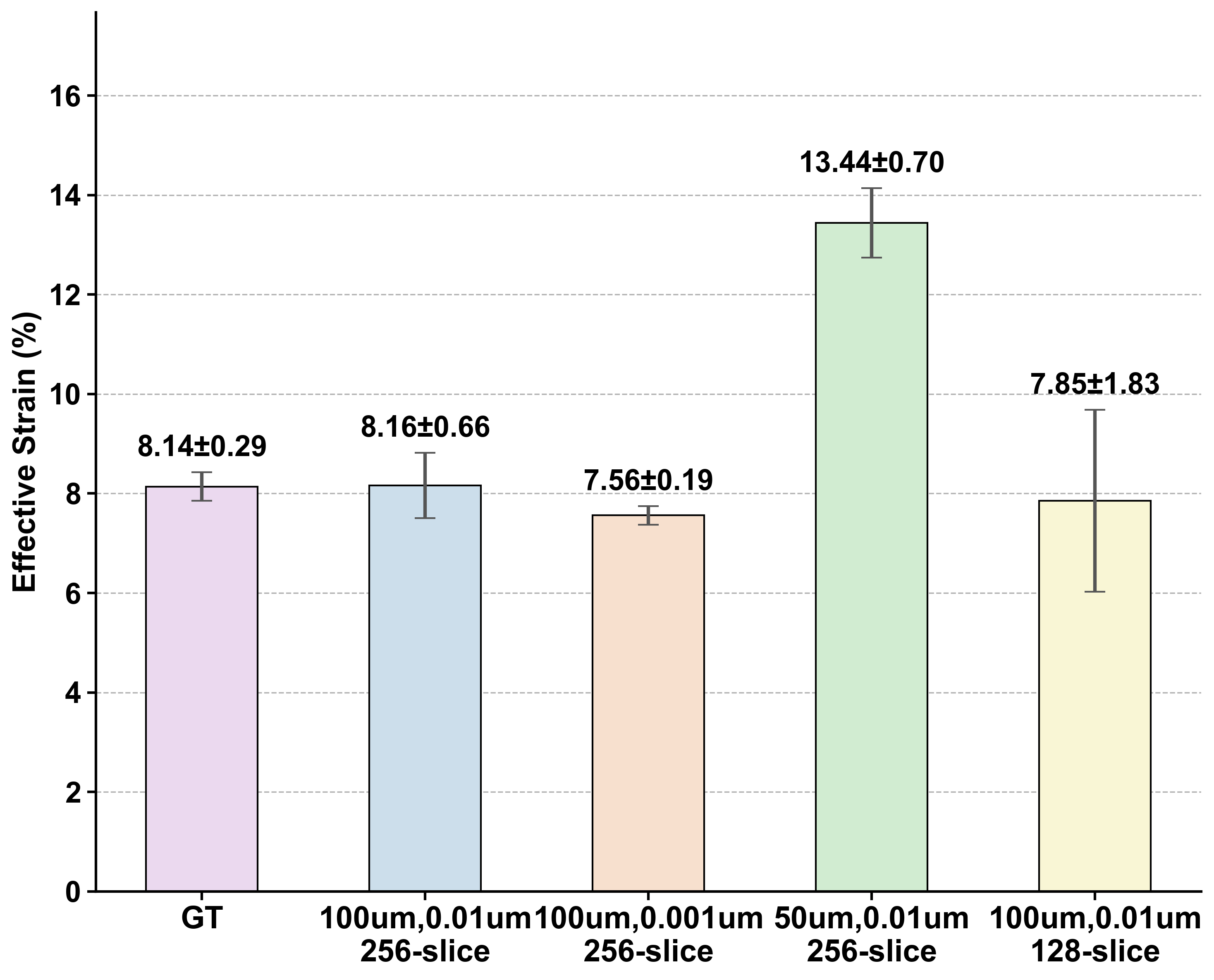

Figure 6. Mean effective strain within the top-20% high-strain region across DVC parameter settings. Bars show mean ± SD across the four validation cases. The x-axis shows GT and DVC settings formatted as mesh size, convergence tolerance, and cropped-volume slice number. GT = ground truth.

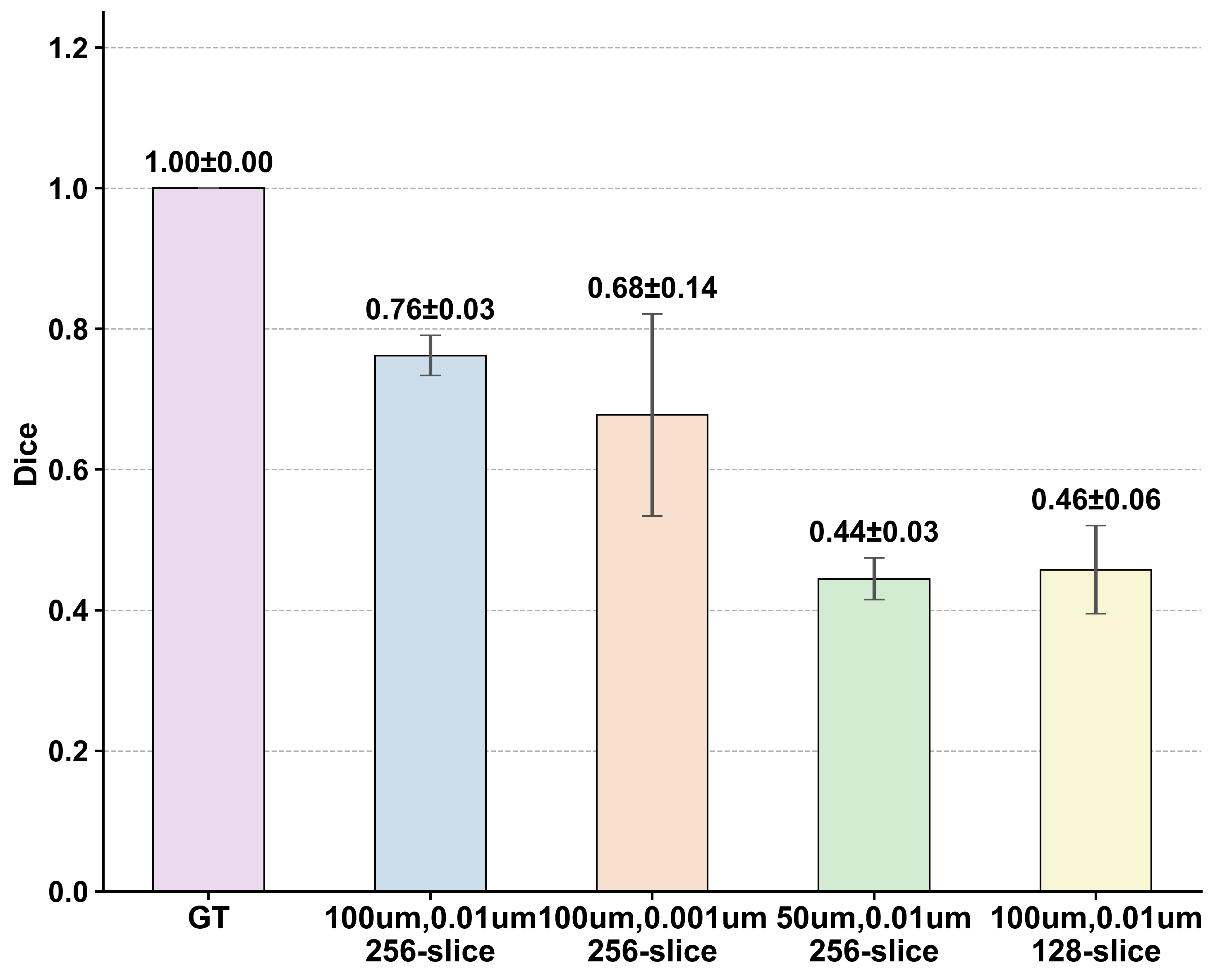

Figure 7. Dice overlap of the top-20% high-strain region between the ground truth and each DVC parameter setting. Bars show mean ± SD across the four validation cases. The x-axis shows GT and DVC settings formatted as mesh size, convergence tolerance, and cropped-volume slice number.

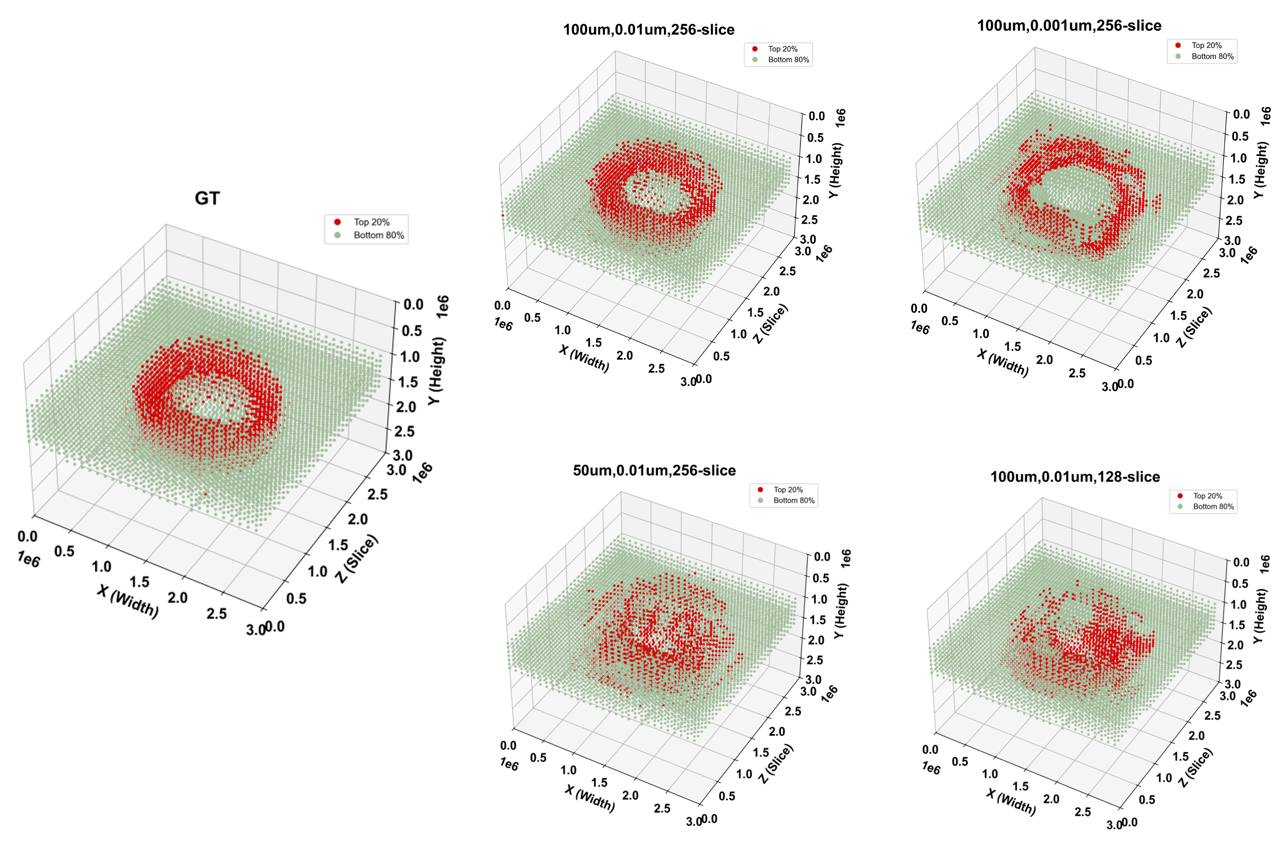

Figure 8. Representative spatial maps of top-20% high-strain regions. Red indicates the high-strain region and green indicates the remaining region, allowing visual comparison between the ground truth and DVC-derived effective-strain maps.
